# Neutralizing antibodies to SARS-CoV-2 Omicron variant after 3^rd^ mRNA vaccination in health care workers and elderly subjects and response to a single dose in previously infected adults

**DOI:** 10.1101/2021.12.22.21268273

**Authors:** Anu Haveri, Anna Solastie, Nina Ekström, Pamela Österlund, Hanna Nohynek, Tuomo Nieminen, Arto A. Palmu, Merit Melin

**Affiliations:** Department of Health Security, Finnish Institute for Health and Welfare, Helsinki, Finland; Department of Information Services, Finnish Institute for Health and Welfare, Helsinki, Finland; Department of Public Health and Welfare, Finnish Institute for Health and Welfare, Tampere, Finland

**Keywords:** SARS-COV-2, neutralizing antibodies, variants of concern, vaccination, COVID-19

## Abstract

The emergence of SARS-CoV-2 Omicron variant (B.1.1.529) with major spike protein mutations has raised concern over potential neutralization escape and breakthrough infections among vaccinated and previously SARS-CoV-2 infected subjects. We measured cross-protective antibodies against variants in health care workers (HCW, n=20) and nursing home residents (n=9) from samples collected 1-2 months following the booster (3rd) dose. We also assessed the antibody responses in prior to Omicron era infected subjects (n=38) with subsequent administration of a single mRNA vaccine dose. Following booster vaccination HCWs had high IgG antibody concentrations to the spike protein and neutralizing antibodies (NAb) were detectable against all variants. IgG concentrations among the elderly remained lower, and some lacked NAbs against the Beta and Omicron variants. NAb titers were significantly reduced against Delta, Beta and Omicron compared to wild-type virus regardless of age. Vaccination induced high IgG concentrations and variable titers of cross-reactive NAbs in previously infected subjects, whereas NAb titers against Omicron were barely detectable 1-month post-infection. High IgG concentrations with cross-protective neutralizing activity were detected after three COVID-19 vaccine doses in HCWs. However, lower NAb titers seen in the frail elderly suggest inadequate protection against Omicron breakthrough infections, yet protection against severe COVID-19 is expected.

**Clinical trial registration:** EudraCT 2021-004788-29

## Introduction

As of 26 November 2021, the World Health Organization (WHO) classified the SARS-CoV-2 B.1.1.529 Omicron variant as a Variant of Concern (VOC) (1). The Omicron variant has been shown to be antigenically more distant from the original SARS-CoV-2 vaccine strain than the previously most distant VOC strains Beta and Delta (2).

**Table 1.**
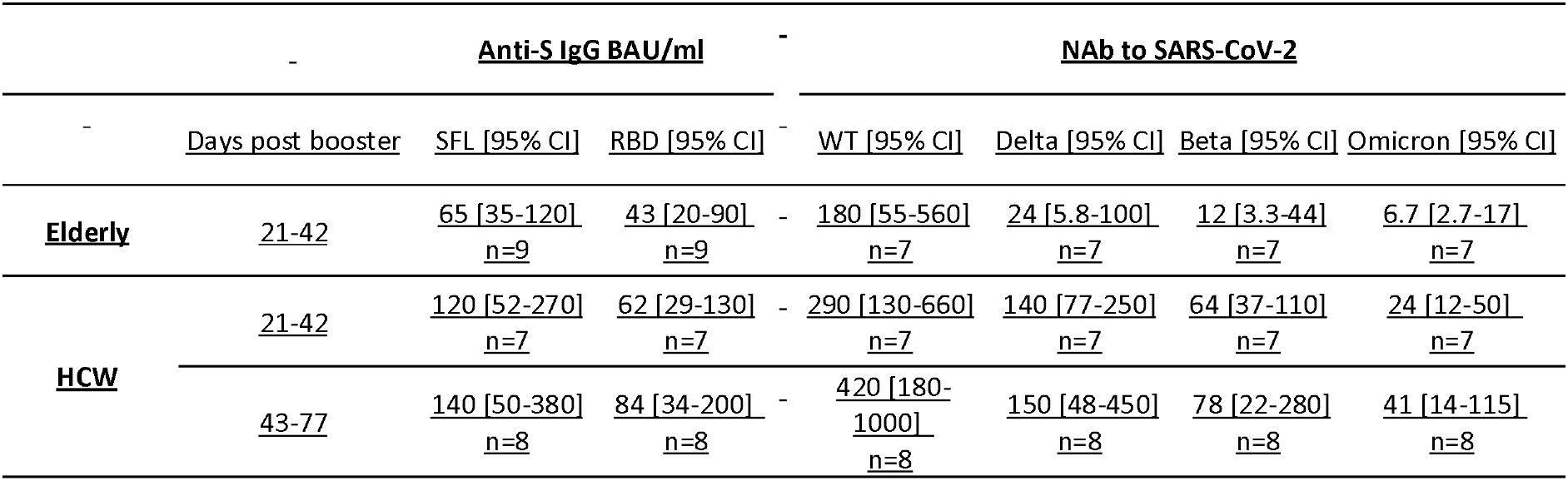
Geometric mean IgG concentrations, GMC [95% CI] expressed as BAU/ml for spike proteins (SFL and RBD) and geometric mean titers, GMT [95% CI] of neutralizing antibodies (NAb) against wild-type (WT) virus and three variants of concern Delta (B.1.617.2), Beta (B.1.351) and Omicron (B.1.1.529) in elderly (n=7-9) and health care workers (HCW) 21-42 (n=7) or 43-77 (n=8) days post booster mRNA vaccination (3^rd^ dose of Comirnaty).

Omicron shares several mutations in the receptor binding domain (RBD) with previously identified variants of concern, including Beta (K417N, N501Y) and Delta (T478K). These mutations influence the ability of the variants to resist the neutralizing activity of antibodies (3-6). Omicron has several additional mutations in the RBD, including E484A (7), that are likely to further enhance its ability to escape neutralizing antibodies (NAbs) and suggest a significant potential for vaccine escape compared with the Delta variant (8).

Omicron has an increased growth rate, household transmission risk and secondary attack rate compared to the Delta variant (9) and it is expected to become the dominant strain worldwide. This has resulted in urgent administration of booster vaccinations and development of variant-specific vaccine compositions.

Both COVID-19 mRNA vaccines were initially shown to be highly immunogenic, also among elderly subjects (10, 11), and were found to be 94 to 95% effective in preventing wild-type (WT) SARS-CoV-2 infection in short-term after the second dose (12, 13). Effectiveness against Delta variant infection has been shown to be slightly lower but remains relatively good at over 80% during the first 3 months, yet lower in the elderly (14). However, longer follow-up data from vaccine effectiveness (VE) studies has indicated that protection wanes more steeply against Delta variant infection. In the elderly, substantial waning to low VE levels of 20% after 5 months has been reported following the second vaccine dose (14). For Omicron variant infection, first VE estimates show considerable and rapid waning after 2 vaccine doses, but restoration of high effectiveness after a mRNA booster (15, 16).

NAb levels are highly predictive of protection against infection and clinical disease (17). However, no absolute antibody titer threshold has been established as a correlate of protection for SARS-CoV-2 (18). Omicron has been shown to escape vaccine-induced humoral immunity; substantially reduced NAb titers have been seen in recently vaccinated subjects (2, 19, 20). Also, substantial ability to evade immunity from prior infection has been suggested (21). However, previously infected subjects with subsequent vaccination were found to retain relatively high neutralization titers (19).

In this study we measured the NAb titers against a WT virus, and three variants isolated in Finland during 2021; Beta (B.1.351), Delta (B.1.617.2) and Omicron (B.1.1.529) in health care workers (HCW) and elderly subjects, who received a booster vaccination 6 to 9 months after the second dose. We also measured NAb responses against Beta, Delta and Omicron as well as Alpha (B.1.1.7) to a single vaccination in prior to Omicron era infected subjects and compared the NAb titers to the peaking antibody response 1 month after infection.

## Results

### Higher IgG antibody concentrations to the SARS-CoV-2 spike protein after booster vaccination in HCW compared to elderly

To estimate the extent of cross-protective immunity following a COVID-19 booster vaccination, we measured the concentration of IgG antibodies to spike protein (anti-S IgG) and NAb titers from HCWs and elderly subjects sampled ≥21 days after the 3^rd^ vaccine dose (Figure 1, Table 1). We found that the anti-S IgG geometric mean concentrations (GMCs) 21 to 42 days following a booster dose (Comirnaty) were 1.4 (RBD) to 1.8-fold (full-length spike glycoprotein, SFL) in HCWs compared to the elderly. The anti-S IgG GMCs were equally high in HCWs sampled at 43 to 77 days after the booster, compared to those sampled earlier. Subjects who received a Spikevax booster had 2.2 (RBD) to 2.5-fold (SFL) anti-S IgG GMCs compared to those who were vaccinated with Comirnaty. None of the differences were statistically significant (Kruskall-Wallis) due to the very small number of subjects.

**Figure 1.**
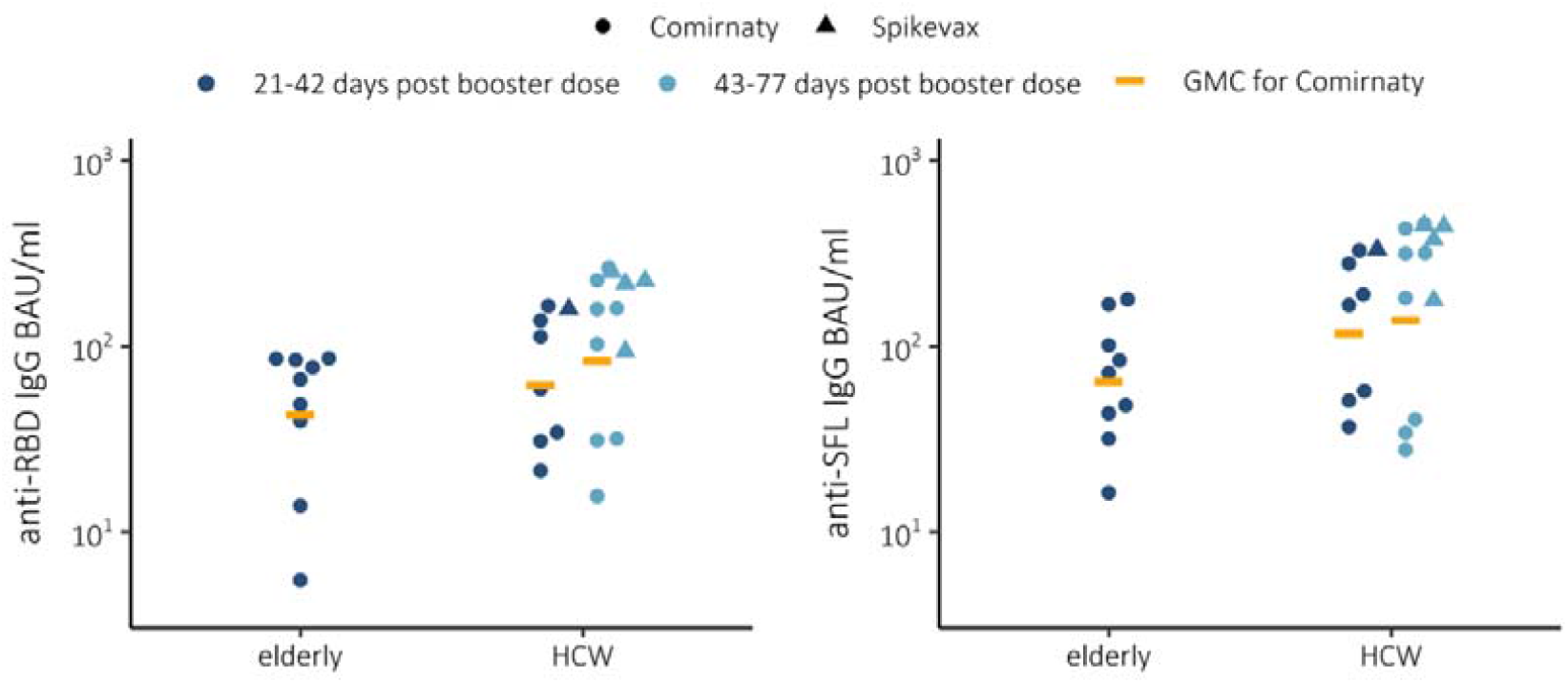
IgG concentrations and geometric mean concentrations (GMC) expressed as BAU/ml for spike proteins (SFL and RBD) in elderly (n=9) and health care workers (HCW) 21-42 (n=8) or 43-77 (n=12) days post booster mRNA vaccination (3rd dose) using Comirnaty. Booster mRNA vaccination Spikevax vaccinated HCWs 21-42 (n=1) and 43-77 (n=4).

### Booster vaccination induced variable concentrations of cross-neutralizing antibodies

Following booster vaccination, we observed measurable NAb titers and likely strong and moderate cross-protection against Delta and Beta variants tested in HCWs, respectively (Figure 2, Table 1). NAb titers to Omicron were decreased by 91% and 96% (*P<*.001 and *P*=.0021, Wilcoxon rank sum test) compared to WT SARS-CoV-2 in HCW and elderly, respectively. NAb titers to Beta variant were decreased by 80% and 93% (*P<*.001 and *P*=.0053) and NAb titers to Delta variant were decreased by 61% and 87% (*P=*.023 and *P=*.029) compared to NAb to WT SARS-CoV-2, in HCWs and elderly, respectively (Figure 3). The NAb titers against WT virus were 1.6-fold, and 3.6-fold against Omicron in HCWs compared to elderly, sampled 21 to 42 days following the Comirnaty booster vaccination (Table 1). The NAb titers against Delta were 4.4-fold, and 3.6-fold compared to Omicron in HCW and elderly, respectively. Furthermore, 2/7 of the elderly did not have NAb to Omicron despite receiving booster dose only 21-42 prior.

**Figure 2.**
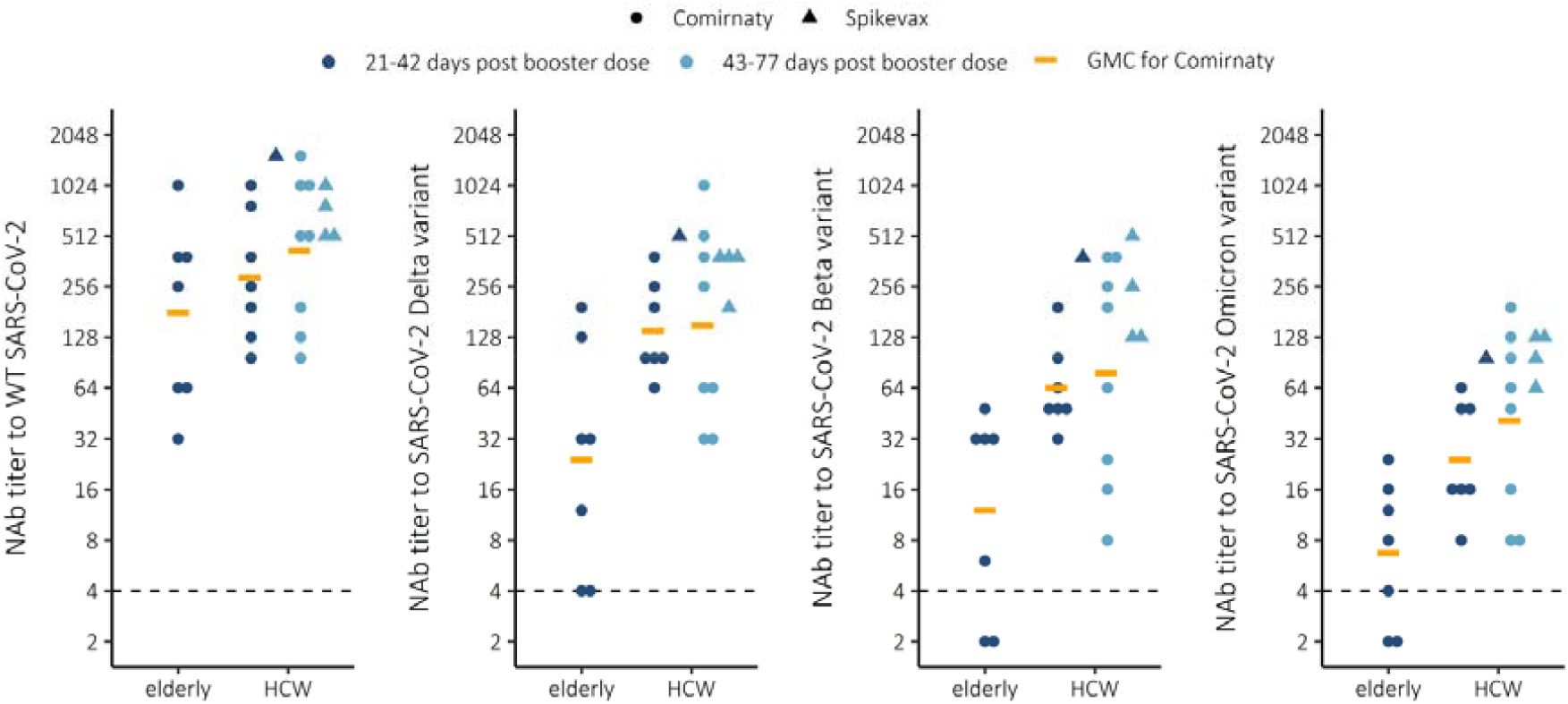
Neutralizing antibody (NAb) titers to wild-type (WT) virus and 3 variants of concern Delta (B.1.617.2), Beta (B.1.351) and Omicron (B.1.1.529) in elderly (n=7) and health care workers (HCW) 21-42 (n=8) or 43-77 (n=12) days post 3rd booster mRNA vaccination with Comirnaty or Spikevax COVID-19 vaccine.

**Figure 3.**
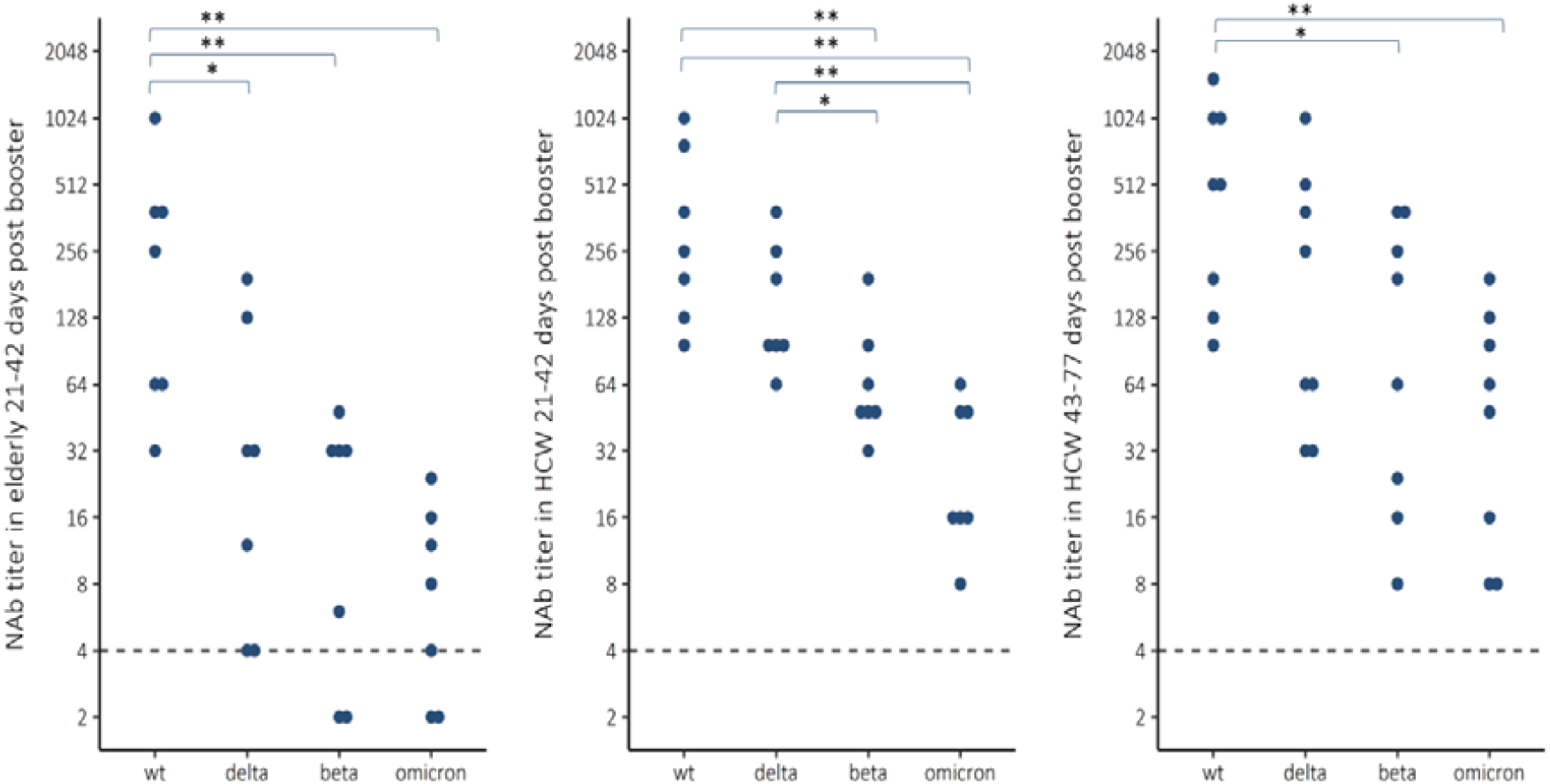
Neutralizing antibody (NAb) titers to wild-type (WT) virus and 3 variants of concern Delta (B.1.617.2), Beta (B.1.351) and Omicron (B.1.1.529) in elderly (n=7) and health care workers (HCW) 21-42 (n=7) or 43-77 (n=8) days post 3rd booster mRNA vaccination with Comirnaty COVID-19 vaccine. Wilcoxon rank sum test *=P<0.05, **=P<0.01.

### A single vaccine dose induced high IgG concentrations and variable NAb titers in previously infected subjects

We measured anti-S IgG concentrations and NAb titers from samples collected from previously infected subjects at 1-2 months following a laboratory-confirmed SARS-CoV-2 infection by WT (n=13), Alpha (n=20) or Beta variant (n=5). The anti-S IgG concentrations measured at 26-70 days after infection were on average 6.3 and 6.4 BAU/ml for RBD and SFL, respectively. The anti-S IgG levels after infection and single dose of Comirnaty COVID-19 vaccine are presented in Supplementary figure S2.

Infection induced strongest NAb titers against the homologous variant strain, but only moderate or low cross-protection against other variants (Figure 4). For Omicron, infection by heterologous variant induced no NAb titers in subjects who had had WT or Beta variant infection, and only 1 subject with prior Alpha variant infection had borderline positive NAb titer against Omicron.

**Figure 4.**
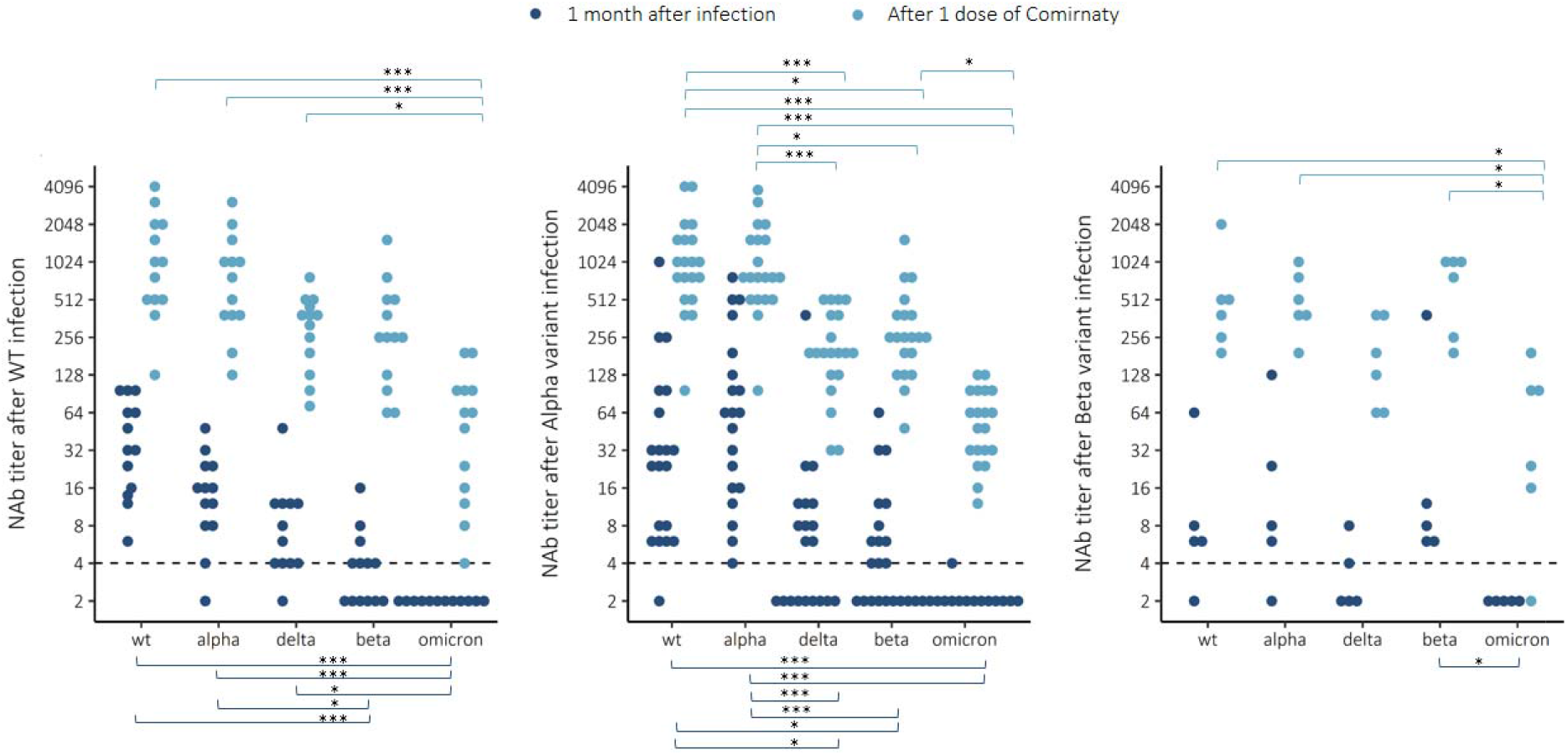
Neutralizing antibody (NAb) titers to wild-type (WT) virus and 4 variants Alpha (B.1.1.7), Delta (B.1.617.2), Beta (B.1.351) and Omicron (B.1.1.529) 1 month after infection with WT, Alpha or Beta variant and after one dose of Comirnaty COVID-19 vaccine. Wilcoxon rank sum test *=P<0.05, **=P<0.01, ***=P<0.001.

We then measured NAb from samples collected from the previously infected subjects who had received a single COVID-19 vaccine dose (Comirnaty) 3 to 6 months following infection. A single dose induced very strong anti-S IgG responses (16 to 27-fold, for RBD and SFL, respectively), and the anti-S IgG concentrations were comparable to the levels following a Comirnaty booster dose in HCWs (75 vs 130 BAU/ml for RBD and 140 vs 170 BAU/ml for SFL) 21-77 days post booster dose. Most importantly, we measured a marked increase in cross-protective antibody responses in previously infected subjects against all variants, inducing Omicron (Figure 4). The NAb titers remained elevated for at least 2 months following vaccination.

## Discussion

Following a booster vaccination, given at 6 to 9 months following the second COVID-19 mRNA vaccine dose, we found high anti-S IgG and detectable NAb concentrations suggesting cross-protection against all variants among HCWs. The anti-S IgG concentrations and the ability to neutralize variants including Omicron remained equally high in samples collected from HCW from 1 to up to 2.5 months following the booster. However, among the frail elderly living in residential care home the antibody concentrations measured a month after the booster dose were lower compared to those among HCW and not all subjects had NAbs against the Beta and Omicron variants.

The age-dependency of the antibody responses suggests that protection against breakthrough infections following a booster dose may be partial and transient in the frail elderly. Antibody responses to COVID-19 vaccination have been shown to correlate negatively with age; vaccination induces highest antibody responses in younger subjects, and lower among subjects aged 65 and older (11, 22, 23). The concentration of NAbs has also been shown to decline faster with increasing age (24, 25).

Furthermore, the response to a booster vaccination is likely dose dependent. The mRNA content in the Comirnaty vaccine is 30 µg compared to 100 µg in the Spikevax; the currently licensed booster dose for Spikevax is 50 µg. A 50 µg Spikevax booster dose was shown to markedly increase the NAb titers against Omicron in subjects previously immunized with two doses of the Spikevax vaccine (26). Preliminary data indicates that a full dose would result in two-fold concentration of neutralizing antibodies against the Omicron variant (27). In this study we found that the HCWs who received a full dose of Spikevax as a booster tended to have higher concentrations of cross-neutralizing antibodies compared to those vaccinated with Comirnaty (2.4-fold against Omicron). However, the difference was not statistically significant due to the small number of subjects.

Effectiveness against infection and severe disease has been reported to be higher for Spikevax compared to Comirnaty (28). In subjects vaccinated with two doses of a mRNA vaccine, Spikevax has been shown to induce two-fold antibody concentrations compared to Comirnaty (23). A high concentration of neutralizing antibodies has previously been shown to correlate with protection against breakthrough infections (29, 30). These data suggest that the stronger the initial immune response to the vaccine, the longer the immunity will persist. Also, our finding that the booster vaccination induced weaker immune responses among the elderly subjects suggests that the added benefit of a booster vaccination against breakthrough infections is likely shorter with older age.

The ability of sera from subjects vaccinated with two doses of a mRNA vaccine (Comirnaty), collected 56 days following the first vaccine dose, to neutralize Omicron variants has been shown to be markedly reduced compared to the ancestral, vaccine-type virus (20). Only 20 and 24% of the vaccinated subjects had NAb against the two Omicron variants tested, compared to 100% against the vaccine-type, Beta and Delta variants (20). Similar findings of very low NAb activity against Omicron were reported in the South-African study of subjects vaccinated with two doses of Comirnaty vaccine (19). Another study using live virus neutralization assay found that subjects recently vaccinated with two doses of an adenovirus vector vaccine (Vaxzevria) or mRNA vaccine (Comirnaty) with an extended interval of 8 to 11 weeks between the two vaccine doses had substantially reduced NAb titers against Omicron, but only some subjects not having any Nab (2). A longer interval between the two doses of the primary series has been shown to result in significantly higher antibody responses indicative of at least partial booster response (31, 32). A booster vaccination, when given as a third dose several months after the second dose, has been shown to significantly increase the IgG and NAb concentrations (32, 33) and markedly improve neutralization of the Omicron variant (26). This is in line with our finding of positive NAb titers against Omicron in all working age subjects, who had received the booster vaccination.

We also found that the anti-S IgG concentrations among previously infected, once-vaccinated subjects were high and NAb titers variable, but comparable to the antibody concentrations following a third vaccine dose among the HCWs. The South-African study also reported that all the previously infected and subsequently vaccinated subjects (5/5) had neutralizing antibodies against Omicron (19). The kinetics of antibody concentrations following infection may be different compared to vaccine-induced immunity, and the breadth of cross-protective immunity may be qualitatively different. Data suggests that although vaccination may induce higher initial antibody concentrations compared to infection, these antibodies appear to decline much faster (34). A follow-up study of 2653 subjects vaccinated with two doses of Comirnaty, and 4361 convalescent COVID-19 patients found that IgG antibody titers decreased by up to 40% each subsequent month in vaccinated subjects, while in the convalescents they decreased by less than 5% per month (34). We have previously shown that anti-S IgG antibodies can be detected up to 13 months following SARS-CoV-2 infection in 97% of the subjects, and that cross-protective NAb against Alpha, Beta and Delta variants persisted in the majority subjects with severe COVID-19 disease (35).

NAb levels induced by infection or vaccination are predictive of protection from symptomatic SARS-CoV-2 infection (17, 29, 30). Although no exact serological correlates have been established, protection from infection likely requires higher NAb levels, whereas the estimated NAb level for protection from severe infection is sixfold lower (Khoury). SARS-CoV-2 infection induces long-lived bone marrow plasma cells that continue to produce antibodies which likely explains why following a rapid decline of antibody concentrations during the first few months, low but stable concentrations can be detected a year after infection (36). Circulating, resting memory B cells have also been detected in convalescent individuals (36); a memory B-cell response to SARS-CoV-2 evolves during the first 6 months after infection, characterized with greater somatic hypermutation, resistance to mutations in the receptor binding domain of the spike protein, and increased potency (37). It is likely that persistent antigen stimulation in previously infected subjects drives this type of maturation of the B cell response. In vaccinated subjects, a booster dose is expected to induce a memory B cell response. In previously infected subjects, vaccination with a single dose of a mRNA vaccine induced not only higher NAb concentrations compared to naïve subjects, but also higher affinity of antibodies, suggesting that in convalescent individuals, the first vaccination recalls pre-existing memory B cells against SARS-CoV-2 that undergo rapid affinity maturation (38). Higher antibody affinity was found to correlate with improved NAb titers against multiple SARS-CoV-2 variants (38). Susceptibility to infection increases over time, as the level of NAbs declines, especially against antigenically different strains such as Omicron (16). However, boosting of immunity with 3^rd^ doses not only increases the concentration of antibodies, but also improves the quality of the B cell response, and potential long-term immunity against severe COVID-19 disease.

We acknowledge several limitations in our study due to the small number of subjects and HCW gender distribution. Also, in HCW and elderly subjects, we analyzed sera after the 3rd dose, but we did not have pre-booster samples available for comparison. However, pre-booster samples likely had very low NAb titers as low Nab activity against Omicron has been reported following the second dose (19, 20, 26); and especially as 6 to 9 months had passed before the 3^rd^ dose was administered. In this study we only assessed humoral immune responses that are critical immune mechanisms reducing infection. SARS-CoV-2 infection and vaccination also induce durable T-cell immunity, which targets multiple epitopes in the SARS-CoV-2 spike protein and is likely to enhance protection against severe COVID-19 disease against SARS-CoV-2 variants (39). Analysis of the mutations in the spike protein sequence of the Omicron variant with computational modelling suggested that the T cell response against Omicron would remain broadly cross-protective, as only a small number of the CD4+ and CD8+ T cell epitopes, and none of the immunodominant epitopes, are affected by the Omicron specific mutations (40).

In summary, the results of our study support previous findings indicating that COVID-19 booster vaccinations raise IgG concentrations, although NAb titers remain low against Omicron compared to WT and Delta variants. In this study we were able to demonstrate measurable NAbs against Delta, Beta and Omicron VOCs up to 2.5 months after 3^rd^ mRNA vaccine dose in adults, whereas some elderly subjects did not have neutralization capacity against Beta and Omicron variants at 1 month after 3^rd^ dose. We observed that the NAb titers against Omicron were barely detectable already 1 month post mild SARS-CoV-2 infection caused by WT virus, Alpha or Beta variant, suggesting an increased risk of reinfection caused by Omicron. However, single mRNA COVID-19 vaccine dose after SARS-CoV-2 infection induced NAb titers comparable to 3^rd^ mRNA COVID-19 vaccination. Booster vaccinations likely improve protection against infections at least temporarily, which may help reduce transmission of the virus in the pandemic situation. Immunity against severe disease is also based on memory B cells and T-cell-mediated immunity, which is expected to improve and last longer after booster vaccination.

## Materials and methods

### Study design and participants

This was an observational clinical vaccine study in which the vaccinations were administered according to the national COVID-19 vaccination campaign.

We invited HCWs (n=20, median age 50.2 [27.2-63.1], 100% female) and elderly in residential care home (n=9, 84.2 [71.5-89.6], 44% female) to participate and donate a blood sample for measurement of SARS-CoV-2 specific serum antibodies (Supplemental Table S1). The HCWs and care home residents had received the 1^st^ and 2^nd^ vaccination (COVID-19 mRNA vaccine, Comirnaty) at a median of 21 [21-35] days apart, and the booster dose (Comirnaty, n=15; Spikevax, n=5) had been given at a median of 245 [206-291] days following the 2^nd^ dose. The blood samples were collected at median of 40 [21-77] days following the booster dose. None of the participants had documentation of a previous SARS-CoV-2 infection in the National Infectious Diseases Register.

Secondly, we invited 480 subjects with laboratory-confirmed SARS-CoV-2 infection identified in the National Infectious Disease Register (between Oct 2020 and March 2021). A total of 82 (17%) subjects participated from which a total of 38 participants with a blood sample taken both before and after their first COVID-19 mRNA vaccine dose (Comirnaty) were selected to this study. The cause of infection of the selected participants had been identified as WT (n=13, median age 54.5 [44.7-80.8], 62% female), Alpha (n= 20, median age 51.2 [27.4-71.4], 55% female) or Beta variant of SARS-CoV-2 (n=5, median age 44.0, [32.7-50.5], 40% female). Only one of the 38 selected subjects had history of severe COVID-19 disease (hospitalization), caused by WT. Blood samples were collected at a median of 50 [26-70] days after the COVID-19 infection and 10 to 20 (n=11), 21 to 42 (n=17) and ≥43 days (n=11) after subsequent administration of a single dose of a COVID-19 mRNA vaccine given at a median of 181, 182, and 158 days after infection, subsequently. From one participant, with Beta variant infection, two post-vaccination samples were included (14 and 76 days after vaccination). COVID19 vaccination history of all the participants was collected from the National Vaccination Registry.

Serum specimens were separated by centrifugation, aliquoted and stored at -20°C or below. For assessment of NAbs, sera were heat-inactivated at 56°C for 30 min.

### SARS-CoV-2 viruses selected for microneutralization test (MNT)

We utilized five SARS-CoV-2 viruses for determining NAb titers. WT virus (B lineage) indicates hCoV-19/Finland/1/2020 (GISAID accession ID EPI_ISL_407079; GenBank accession ID MZ934691). WT virus isolation and propagation were performed in African green monkey kidney epithelial (Vero E6) cells (41). All variant viruses were isolated and propagated in VeroE6-TMPRSS2-H10 cells (42) and further propagated in Vero E6 cells for MNT. Alpha variant (B.1.1.7) indicates the isolate hCoV-19/Finland/THL-202102301/2021 (EPI_ISL_2590786; MZ944886), Beta variant (B.1.351) the isolate hCoV-19/Finland/THL-202101018/2021 (EPI_ISL_3471851; MZ944846), Delta variant (B.1.617.2) the isolate hCoV-19/Finland/THL-202117309/2021 (EPI_ISL_2557176; MZ945494) and Omicron variant (B.1.1.529) the isolate hCoV-19/Finland/THL-202126660/2021 (pending for the GISAID and GenBank ID).

### SARS-CoV-2 MNT

We performed a cytopathic effect-based MNT as previously described (35, 41). Briefly, serum samples were 2-fold serially diluted starting from 1:4 in Eagle’s minimum essential medium supplemented with penicillin, streptomycin and 2% of heat-inactivated fetal bovine serum. At the biosafety level 3 laboratory, pre-titrated virus was added to obtain 100 x tissue culture infectious dose 50% per well following incubation for 1 h at +37°C, 5% CO2. African green monkey kidney epithelial (VeroE6) cells were added, and the 96-well tissue culture plates were incubated at +37°C, 5% CO2 for 4 days. Wells were fixed with 30% formaldehyde and stained with crystal violet. Results were expressed as MNT titers corresponding to the reciprocal of the serum dilution that inhibited 50% of SARS-CoV-2 infection observed by the cytopathic effect of inoculated cells. MNT titer ≥6 was considered positive, borderline when 4 and negative when <4. Borderline values were further confirmed with biological repeats. For titer comparison, a titer of 192, 8, 32 and <8 was measured for the WHO International Standard (NIBSC 20/136 (43)) using the WT virus, Beta, Delta and Omicron variant, respectively.

### SARS-CoV-2 fluorescent multiplex immunoassay

We measured the concentration of IgG antibodies to spike glycoprotein of SARS-CoV-2 (S-IgG Ab) with in-house fluorescent multiplex immunoassay as previously described (44). Briefly, sera from donors were incubated with microspheres covered with SARS-CoV-2 receptor binding domain and full-length spike glycoprotein, and bound antibodies were detected with R-Phycoerythrin (RPE)-conjugated secondary antibody. Antibody concentrations were measured with MAGPIX^®^ system using xPONENT software v4.2 (Luminex^®^Corporation, Austin, TX) and converted into BAU/ml, which is calibrated with WHO international standard for SARS-CoV-2 antibody assays.

### Statistical methods

The study analysis is descriptive. We calculated the geometric mean concentrations (GMC) and titers GMTs with 95% confidence intervals (CI) for IgG and NAb levels, respectively. MNT titers <4 were assigned a titer value of 2. We assessed the statistical differences in antibody levels between groups using the Kruskal-Wallis test with Bonferroni correction. Differences in neutralizing antibody titers between viruses were assessed with Wilcoxon rank sum test. The statistical significance level of difference was set to p<0.05. Statistical analyses were performed using SPSS v27 and R (v4.0.4) with Rstudio (v1.4.1106).

## Data Availability

Data subject to third party restrictions. The data that support the findings of this study are available from Finnish Social and Health Data Permit Authority Findata. Restrictions apply to the availability of these data, which were used under license for this study under informed consent form. Anonymized data are available after permission by Findata.

## Ethics approval and patient consent statement

The study protocol of the COVID-19 vaccine immunological studies in Finland was approved by the National Committee on Medical Research Ethics (TUKIJA/347/2021) and by the Finnish Medicines Agency Fimea as the regulatory authority (European Union clinical trials database code of EudraCT 2021-004788-29). For the follow-up of COVID-19, the study protocol of the serological population study of the coronavirus epidemic was approved by the ethical committee of the Hospital District of Helsinki and Uusimaa (HUS/1137/2020). Written informed consent was obtained from all study subjects before sample collection.

## Author contributions

M.M. and A.H. designed the experiments. M.M., A.A.P. and H.N. contributed to the study design. A.S. performed the FMIA tests. A.H. developed and performed the microneutralization tests. P.Ö. coordinated the virus isolations. A.S., A.H. and T.N. analyzed the data. N.E coordinated the participant recruitment, sample collection and sample processing. A.H., A.S., N.E. and M.M. wrote the manuscript and all co-authors contributed to the critical revision of the text.

## Acknowledgements

We thank all the study participants. We also thank Juha Oksanen, Esa Ruokokoski, Elina Isosaari, Tommi Korhonen, Joni Niemi, Oona Liedes, Saimi Vara, Päivi Siren, Maila Kyrölä, Ritva Syrjänen, Heta Nieminen, Camilla Virta, Lotta Hagberg, Marja Leinonen, Arja Rytkönen, Mervi Lasander, Marja-Liisa Ollonen, Larissa Laine, Marika Skön, Tiina Sihvonen, Elina Virtanen, Johanna Rintamäki, Leena Saarinen, Marja Suorsa, Minna Haanpää, Johanna Mustajoki, Mervi Eskelinen, Niina Ikonen, Kirsi Liitsola, Soile Blomqvist, Erika Lindh and Esa Rönkkö.

We gratefully acknowledge the authors and their respective laboratories, who analyzed and submitted the sequences to GISAID’s EpiCoV™ and GenBank^®^ Database.

This study was funded by the Finnish institute for Health and Welfare and the Academy of Finland (Decision number 336431).

## Abbreviations

BAU: Binding antibody unit concentration
COVID-19: Coronavirus Disease 2019
GMC: Geometric mean concentration
GMT: Geometric mean titer
HCW: Health care worker
MNT: Microneutralization test
mRNA: Messenger ribonucleic acid
NAb: Neutralizing antibody
RBD: Receptor-binding domain
SARS-CoV-2: Severe acute respiratory syndrome coronavirus 2
SFL: Full length spike glycoprotein
anti-S IgG: IgG antibodies to spike protein
VE: Vaccine effectiveness
VOC: Variants of concern
WT: Wild-type

