## Supplemental Figure 1 for "Neutralizing antibodies to SARS-CoV-2 Omicron variant after 3^rd^ mRNA vaccination in health care workers and elderly subjects and response to a single dose in previously infected adults"

**Supplementary Figure S1.** Distribution of days between sampling and mRNA booster dose and it’s effect on neutralizing antibody (NAb) titers to wild-type (WT) virus and 3 variants of concern Beta (B.1.351), Delta (B.1.617.2) and Omicron (B.1.1.529) in elderly (n=7) and health care workers (HCW) (n=20).

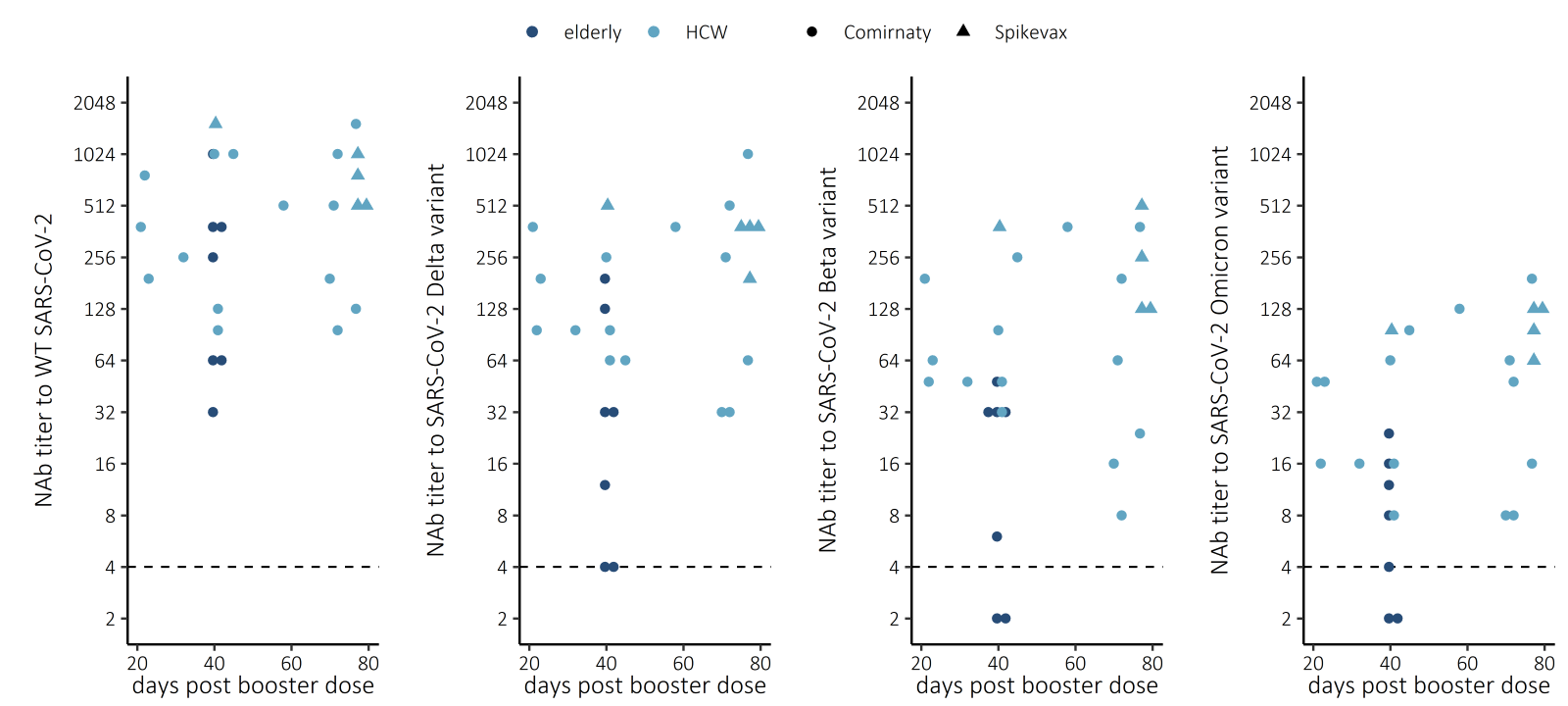
