## Supplemental Figure 2 for "Neutralizing antibodies to SARS-CoV-2 Omicron variant after 3^rd^ mRNA vaccination in health care workers and elderly subjects and response to a single dose in previously infected adults"

**Supplementary Figure S2.** IgG concentrations expressed as BAU/ml for spike proteins (SFL and RBD) 1 month after infection with WT, Alpha or Beta variant and after 1 dose of Comirnaty COVID-19 vaccine.

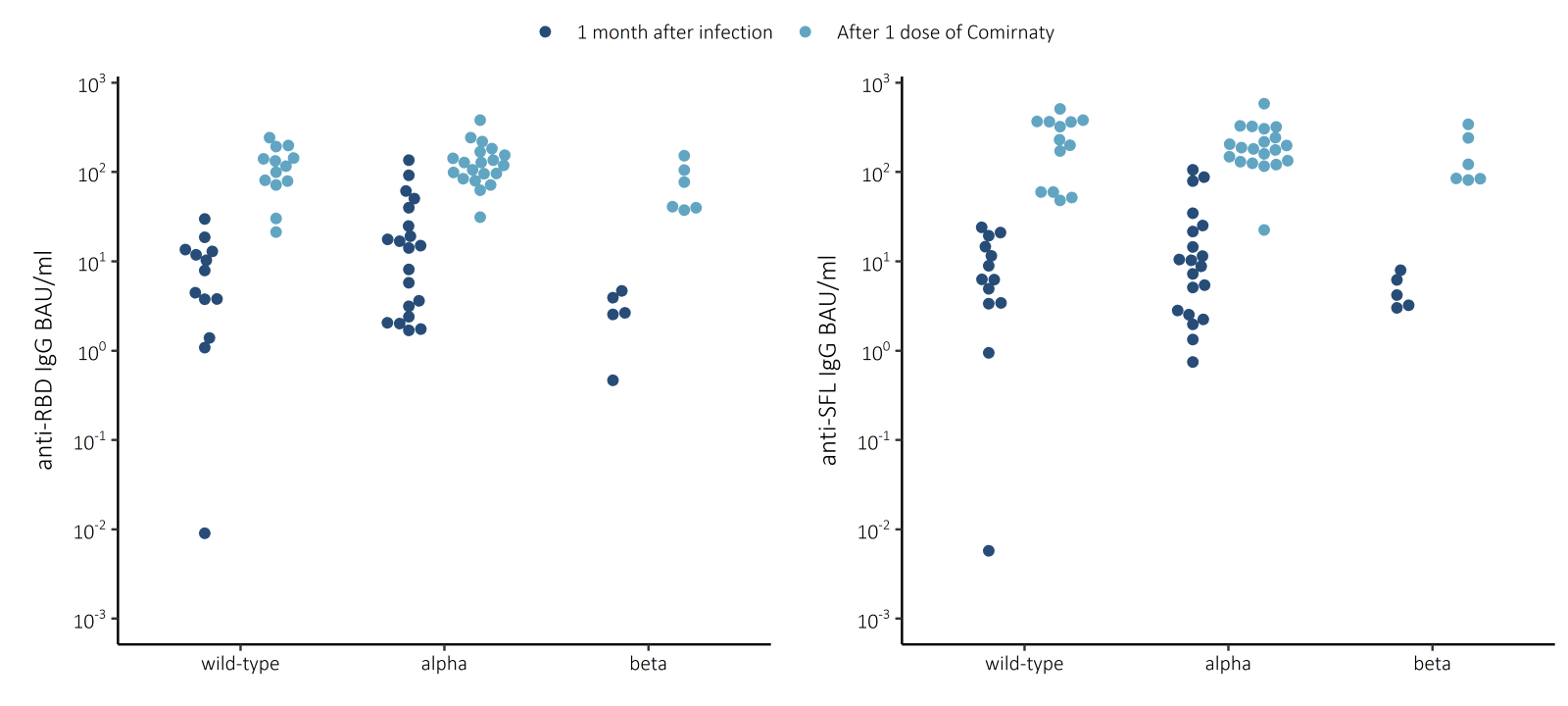
