## Supplemental Table 1 for "Neutralizing antibodies to SARS-CoV-2 Omicron variant after 3^rd^ mRNA vaccination in health care workers and elderly subjects and response to a single dose in previously infected adults"

**Supplementary Table S1.** Demographic characteristics of the participants.

|  | **Age**  **Median [range]** | **Gender**  **% female** |
| --- | --- | --- |
| Health care workers (n=20) | 50.2 [27.2-63.1] | 100 |
| Elderly  (n=9) | 84.2 [71.5-89.6] | 44 |
| Infected (Wild-type)  (n=13) | 54.5 [44.7-80.8] | 62 |
| Infected (Alpha)  (n=20) | 51.2 [27.4-71.4] | 55 |
| Infected (Beta)  (n=5) | 44.0 [32.7-50.5] | 40 |
